# High-dimensional immune profiling of dimethyl fumarate and ocrelizumab in multiple sclerosis

**DOI:** 10.1101/2022.11.05.22281806

**Authors:** Yinan Zhang, Brian Lee, Hui Xie, Jonathan Rockoff, Sammita Satyanarayan, Rachel Brandstadter, Seunghee Kim-Schulze, Achillefs Ntranos, Fred Lublin

**Affiliations:** Department of Neurology, The Ohio State University Wexner Medical Center, Columbus, OH; Human Immune Monitoring Center, Icahn School of Medicine at Mount Sinai, New York, NY; Department of Neurology, Icahn School of Medicine at Mount Sinai, New York, NY; Department of Neurology, University of Pennsylvania Perelman School of Medicine, Philadelphia, PA

**Keywords:** multiple sclerosis, dimethyl fumarate, ocrelizumab, lymphocyte subsets, biomarkers, proteomics

## Abstract

**Background:** Dimethyl fumarate (DMF) and ocrelizumab are two effective immunomodulators for multiple sclerosis (MS) with distinct mechanisms of action. Identifying overlapping therapeutic effects between both agents may elucidate common pathways in preventing disease activity.

**Objectives:** In this study we analyzed cytokine and immune-profiling data to evaluate the similarities and differences between the two disease-modifying therapies for MS. *Methods*: Plasma and PBMCs from MS patients were collected at baseline, 3 months and 6 months after treatment with DMF (n=16) and ocrelizumab (n=13). Immunophenotyping was performed with mass cytometry (CyTOF) and analyzed with gating based on cell surface markers. Cytokine analysis from plasma was performed with Olink assays and analyzed with linear mixed effects models.

**Results:** DMF reduced both effector T and memory B cell populations while increasing CD56^bright^ natural killer (NK) cells. Ocrelizumab exerted its main immunomodulatory effect by reducing the frequency of all B cells and increasing frequency of NK cells. At 6 months, naive B-cells began to reconstitute; however, memory B cells remain depleted. DMF treatment was associated with a significant reduction of plasma cytokines involved in inflammatory pathways, such as IL-6, IL-12, and Dectin-1 signaling. In addition, DMF lowered plasma cytokines that are dysregulated in psoriasis and involved in allograft rejection pathways. Ocrelizumab treatment led to the upregulation of neurotropic proteins in the plasma of MS patients, including proteins involved in NAD+ biosynthesis and tryptophan catabolism.

**Conclusion:** Our high-dimensional immunophenotyping results suggest that to exert their effects on MS patients, DMF and ocrelizumab both increase NK cells in addition to affecting different immune cell populations and cytokine pathways. Detecting similarities between the mechanisms of the two drugs may contribute to identifying more specific therapeutic targets.

## Introduction

Disease-modifying therapies (DMT) for multiple sclerosis (MS) suppress inflammatory activity through different mechanisms of action. Dimethyl fumarate (DMF) and ocrelizumab are two approved therapies for MS. DMF is a twice daily oral DMT. Its metabolite, monomethyl fumarate (MMF), exerts therapeutic effects largely through downregulation of immune responses by activating the Nrf2-anti-oxidative pathway and suppressing the NFkβ inflammatory pathway.^1^ Previous studies have shown DMF depletes lymphocytes in the adaptive immune system including memory B cells and memory CD4 and CD8 T cells while increasing naive and transitional cells.^2-8^ In contrast, ocrelizumab is a monoclonal antibody against CD20, administered as an infusion every 6 months. Treatment with ocrelizumab depletes immature and mature B cells but spares plasma cells and hematopoietic stem cells due to their lack of CD20 expression. In addition to demonstrating efficacy in reducing the frequency of clinical relapses and new MRI lesion formation in relapsing-remitting MS (RRMS), ocrelizumab has been shown to be effective in inhibiting disability progression in primary progressive MS (PPMS).^9^

Despite increasing understanding of the immunophenotype changes with DMF and ocrelizumab therapy, the mechanisms by which these alterations of immune profiles modify effector function is not fully understood. In this study we conducted high-dimensional immune profiling of MS patients treated with DMF or ocrelizumab to characterize changes in lymphocyte composition and proteomic biomarkers. In addition, we performed gene set enrichment analysis to identify pathways affected by treatment.

## Methods

### Study participants

The study enrolled 29 adult patients with either clinically-isolated syndrome (CIS), RRMS, secondary progressive MS (SPMS), or PPMS from the Corinne Goldsmith Dickinson Center for Multiple Sclerosis at Mount Sinai. Patients were either treatment-naive or had at least 90 days washout period from a previous DMT. Participants also have not received steroids or antibiotics in the past 30 days. Blood samples were obtained before therapy with DMF or ocrelizumab and at 3 and 6 months after starting treatment. Informed consent was obtained from all patients prior to participation in the study, which received ethical approval by the Mount Sinai institutional review board. Patients were followed clinically for the duration of the study under standard outpatient care for MS. Clinical outcomes including relapses, Expanded Disability Status Scale (EDSS) score, and MRI data were also collected.

### Mass cytometry

PBMCs were incubated for 20 minutes in a 37LC water bath with Cell-ID Rh103 Intercalator (Fluidigm, Cat# 201103A) in cell culture media to label non-viable cells. Timepoint samples from the same patient were then barcoded with CD298+B2M antibodies conjugated to a unique metal isotope, washed twice and pooled for each patient to minimize staining variability within patient timepoint samples. The pooled patient samples were then blocked with Fc receptor blocking solution (Biolegend, Cat# 422302) and stained with a cocktail of surface antibodies for 30 minutes on ice. All antibodies were either obtained commercially from Fluidigm or conjugated in-house using Fluidigm’s X8 polymer conjugation kits. Pooled-timepoint patient samples were then fixed and barcoded with Fluidigm’s 20-Plex barcoding kit (Cat# 201060) and pooled into a single tube. The sample was then re-fixed with 2.4% paraformaldehyde in PBS containing 0.02% saponin and Cell-ID Intercalator-Ir (Fluidigm, Cat# 201192A) to label nucleated cells. The cells were then stored as a pellet in PBS until acquisition.

Immediately prior to acquisition, the pooled sample was washed with Cell Staining Buffer and Cell Acquisition Solution (Fluidigm, Cat# 201240) and resuspended in Cell Acquisition Solution at a concentration of 1 million cells per ml containing a 1:20 dilution of EQ normalization beads (Fluidigm, Cat# 201078). The samples were acquired on the Fluidigm Helios Mass Cytometer at an acquisition speed of under 400 cells per second.

The resulting FCS data files were normalized and concatenated using Fluidigm’s CyTOF software and de-multiplexed using the Zunder lab single-cell debarcoder (https://github.com/zunderlab/single-cell-debarcoder). The FCS files were further cleaned on Cytobank by removing EQ beads, low DNA debris and gaussian multiplets. Barcoding multiplets were also removed based on the mahalanobis distance and barcode separation distance parameters provided by the Zunder lab debarcoder.

The cleaned FCS files were then uploaded to the Astrolabe Diagnostics platform for automated cell annotation and downstream analysis.

### Proteomic analysis

Serum protein levels were measured using a 96-plex immunoassay from Olink’s Inflammation and Neurology panels.^10^ The Inflammation panel includes 92 inflammatory biomarkers, composed predominantly of cytokines and chemokines associated with inflammatory diseases and related biological processes. The Neurology panel features 92 neurology-related protein biomarkers related to neurological diseases in addition to proteins with roles in cellular regulation, immunology, development, and metabolism.

The assay utilizes proximity extension technology, in which oligonucleotide-tagged antibodies binding to epitopes causes hybridization of matched tags that are then amplified using quantitative PCR and read in log base-2 normalized protein expression values.

### Pathway enrichment analysis

Gene ontology pathway enrichment was performed using Enrichr (https://amp.pharm.mssm.edu/Enrichr), a web-based enrichment tool where gene set inputs are checked for significant overlaps with annotated gene sets from extensive libraries to identify a list of enrichment terms such as pathways and cell lines.^11^ The input gene sets for the enrichment analysis consisted of genes encoding proteins showing significant change (p<0.10) for at 6 months compared to baseline for treatment with DMF or ocrelizumab. The threshold value of enrichment was set at p<0.10. The enriched pathways were displayed on the webpage and downloaded as images.

### Statistical analysis

Bivariate gating was used to define and enumerate immune populations following gating strategy developed by Cytobank (Santa Clara, CA). Results indicate paired analysis with Wilcoxon signed-rank test with FDR correction applied for multiple comparisons. Probability values of <0.05 were considered significant. For cytokine analysis, both Olink Immunology and Neurology panels were merged into a single dataset and analyzed in pairs with the FDR corrected Wilcoxon signed-rank test. All statistical analyses were performed in R.

## Results

### Participant demographics and clinical data

Of the 29 participants enrolled in the study, 13 received DMF and 16 were treated with ocrelizumab. All patients starting DMF were naive to DMT treatment. The majority (n=10) of patients starting ocrelizumab were treatment naive and the rest received prior treatment with either interferon beta, glatiramer acetate, teriflunomide, DMF, or natalizumab with a 90-day minimum washout period. Patients treated with DMF in general did not have any new disease activity; 1 patient had a clinical relapse and 5 patients had new MRI lesions during the study period. No patients receiving ocrelizumab had a clinical relapse or new MRI lesions during the study period. Table 1 summarizes patient demographics and clinical data.

**Table 1.** Patient demographics and clinical data during the 6-month study period. CIS; clinically-isolated syndrome; RRMS: relapsing-remitting multiple sclerosis (MS); SPMS: secondary progressive MS; PPMS: primary progressive MS; EDSS: expanded disability status scale.

| **Characteristics** | **Dimethyl fumarate**<br>**(n=16)** | **Ocrelizumab (n=13)** |
| --- | --- | --- |
| **Mean age** | 50.2 | 35.4 |
| **Sex** |  |  |
| Female | 5 (38.5%) | 11 (68.8%) |
| Male | 8 (61.5%) | 5 (31.2%) |
| **Disease phenotype** |  |  |
| CIS | 0 (0.0%) | 2 (12.5%) |
| RRMS | 8 (61.5%) | 14 (87.5%) |
| SPMS | 1 (7.7%) | 0 (0.0%) |
| PPMS | 4 (30.8%) | 0 (0.0%) |
| **Median EDSS** | 2.5 | 1.5 |
| **Relapse in past year** |  |  |
| Yes | 7 (53.8%) | 15 (93.8%) |
| No | 6 (46.2%) | 1 (6.2%) |
| **MRI activity in past year** |  |  |
| Yes | 9 (81.8%) | 8 (80.0%) |
| No | 2 (18.2%) | 2 (20.0%) |
| **New relapse** |  |  |
| Month 3 | 0 (0.0%) | 0 (0.0%) |
| Month 6 | 1 (6.3%) | 0 (0.0%) |
| **New MRI activity** |  |  |
| Month 3 | 1 (6.3%) | 0 (0.0%) |
| Month 6 | 4 (23.5%) | 0 (0.0%) |

### DMF reduced effector T and memory B cells and increased CD56^bright^ natural killer cells

We assessed the treatment effect of DMF on the peripheral lymphocyte subsets to confirm prior findings of DMF therapy on immune profile changes. At 6 months, DMF significantly reduced effector CD4 and CD8 T cells (Figure 1). Among CD4+ T cells, proportions of Th1, Th17 and Th1-like Th17 cells were also reduced at 6 months of therapy, but regulatory T cells remained unchanged during the study period. Effects on B lymphocytes were seen in the reduction of memory B cells without changes in total B cells. Populations of NK cells including late NK cells, CD56^bright^ NK cells, as well as immature B cells and naive CD4 cells displayed an opposite trend, showing significant increases with DMF treatment at 6 months.

**Figure 1.**
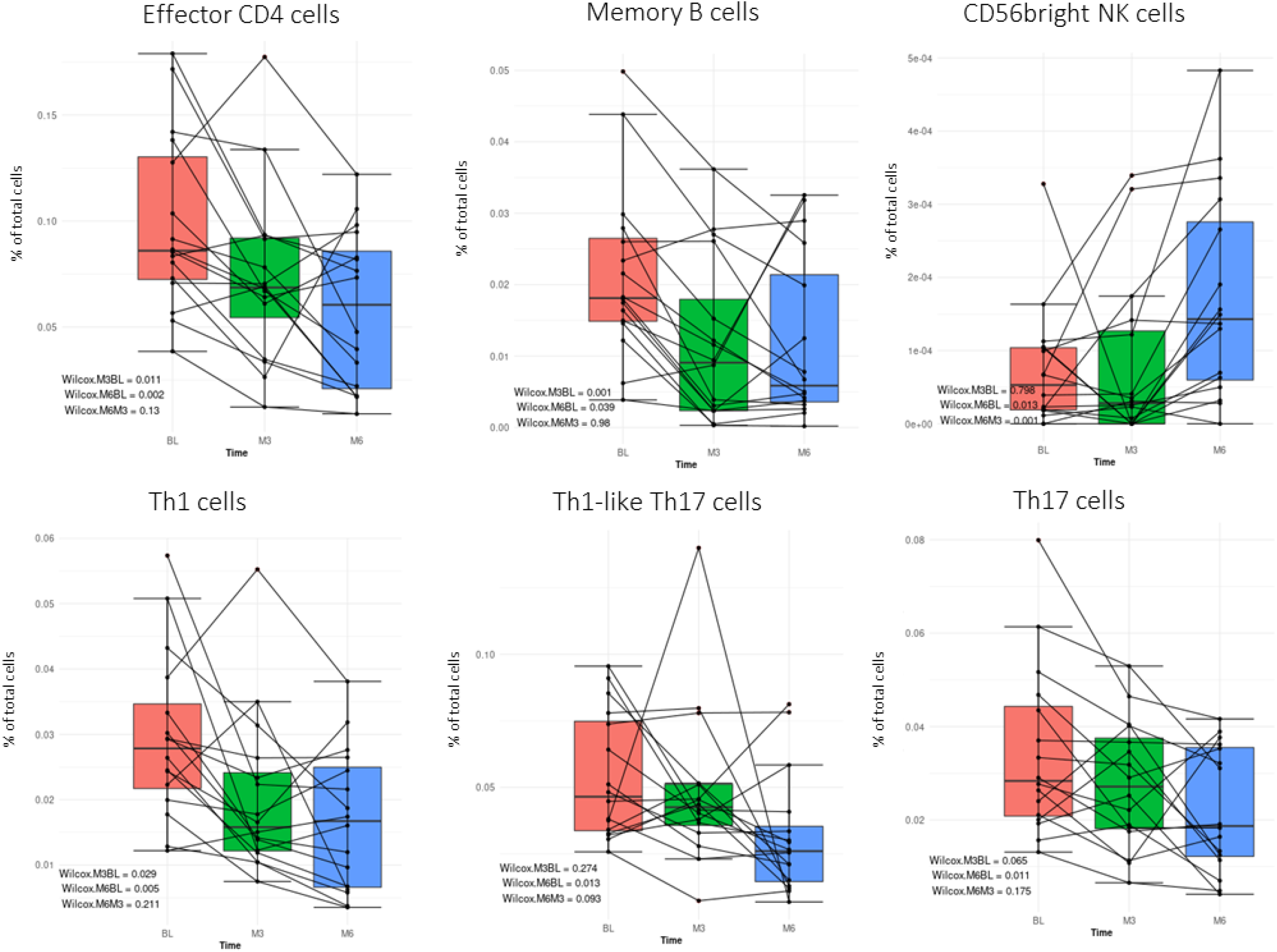
Mass cytometry analysis of immune profiling changes in 16 patients taking dimethyl fumarate (DMF) at baseline (BL), 3 months (M3) and 6 months (M6). Each line represents values for an individual patient at the respective time point in the study. The box plot shows the range (whiskers), first and third quartiles (box), and median (line in box) of the values for each time point. The exact p values shown were calculated using Wilcoxon signed-rank test.

### Ocrelizumab depleted B cells and increased frequency of natural killer cells

As expected of the anti-CD20 effects of ocrelizumab, the main immunomodulatory effect was seen in the robust depletion of all B cells (Figure 2). At 6 months, naive B-cells began to reconstitute; however, memory B cells remain depleted. There was also an increase in the frequency of early and late NK cells and CD56-CD16+ NK cells. There was a trend for reduction of CD3+CD20^dim^ T cells, but this did not reach statistical significance. Total T cells were unchanged, including CD20 positive T cells.

**Figure 2.**
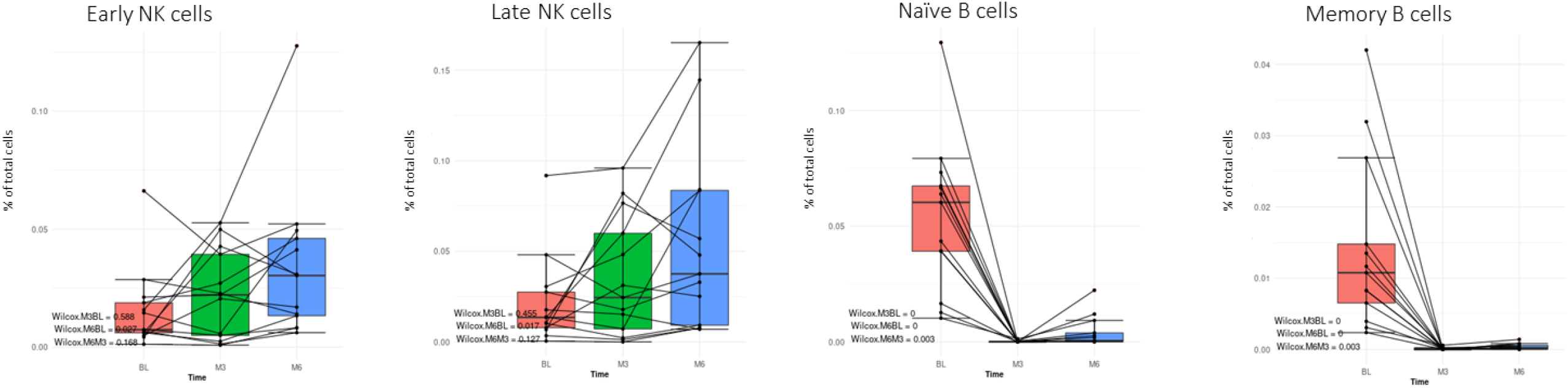
Mass cytometry analysis of immune profiling changes in 13 patients taking ocrelizumab at baseline (BL), 3 months (M3) and 6 months (M6). Each line represents values for an individual patient at the respective time point in the study. The box plot shows the range (whiskers), first and third quartiles (box), and median (line in box) of the values for each time point. The exact p values shown were calculated using Wilcoxon signed-rank test.

### DMF reduced pro-inflammatory cytokines involved in dectin-1 signaling, psoriasis, and IL6/IL12 signaling pathways

DMF treatment was associated with a significant reduction of plasma cytokines involved in inflammatory pathways including CXCL9, IL-12, IL-10, FLT3L, TNFRSF9, CRTAM, SIGLEC1, CDCP1 (Figure 3). Genes coding for these cytokines were uploaded to Enrichr for pathway enrichment analysis. The GO terms revealed that the genes were associated with proteins involved in psoriasis, dectin-1 signaling, and IL6/IL12 signaling pathways (Figure 5).

**Figure 3.**
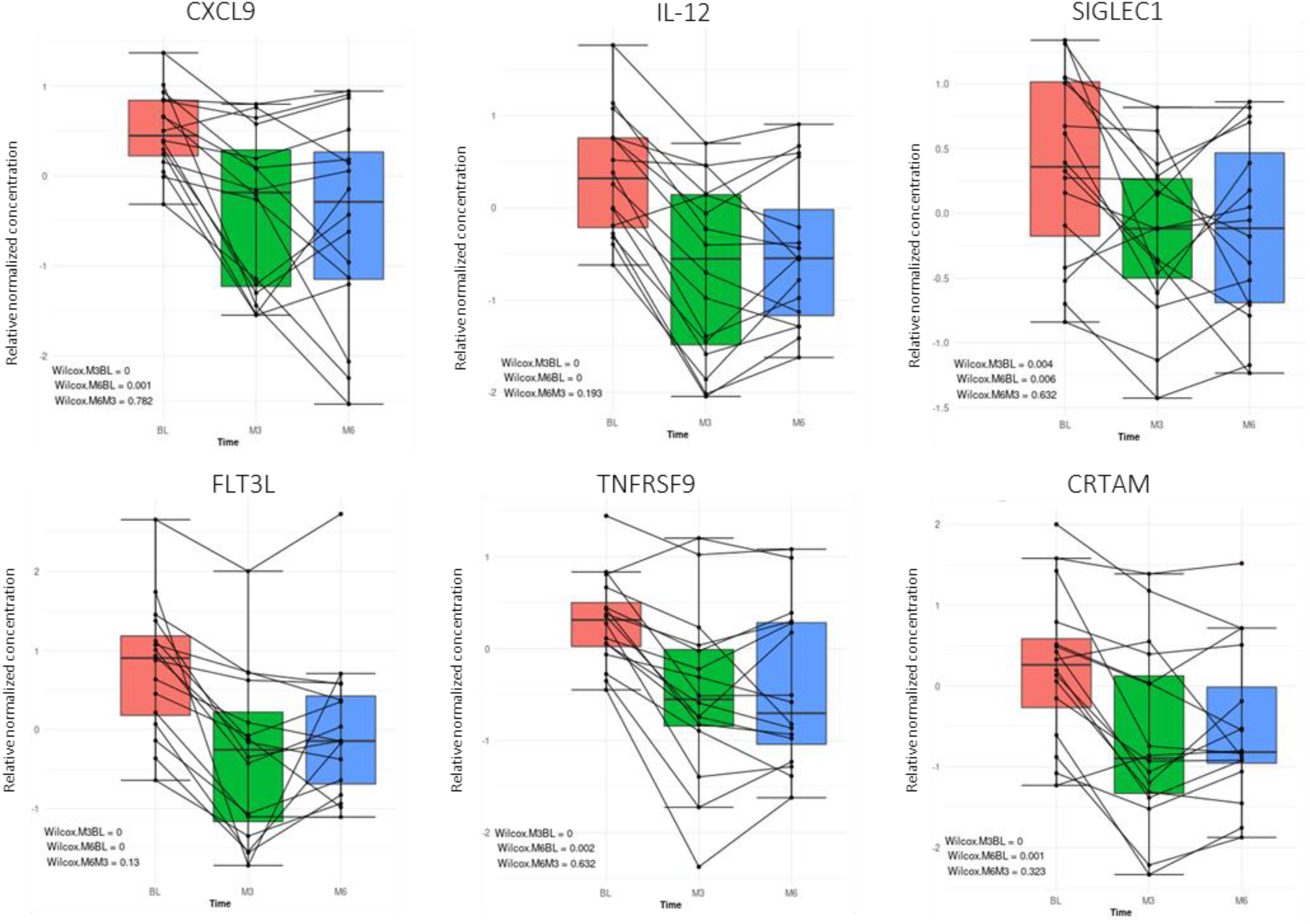
Olink assay analysis of changes in cytokines in 16 patients taking dimethyl fumarate (DMF) at baseline (BL), 3 months (M3) and 6 months (M6). Each line represents values for an individual patient at the respective time point in the study. The box plot shows the range (whiskers), first and third quartiles (box), and median (line in box) of the values for each time point. The exact p values shown were calculated using Wilcoxon signed-rank test.

### Ocrelizumab increased plasma neurotropic proteins including proteins involved in NAD biosynthesis and tryptophan metabolism

Ocrelizumab treatment led to the upregulation of neurotropic proteins in the plasma of MS patients including KYNU, NMNAT1, SPOCK1, FCRL2, GCP5, and NRP2 (Figure 4). Gene set enrichment revealed pathways associated with proteins involved in NAD biosynthesis and tryptophan catabolism (Figure 5).

**Figure 4.**
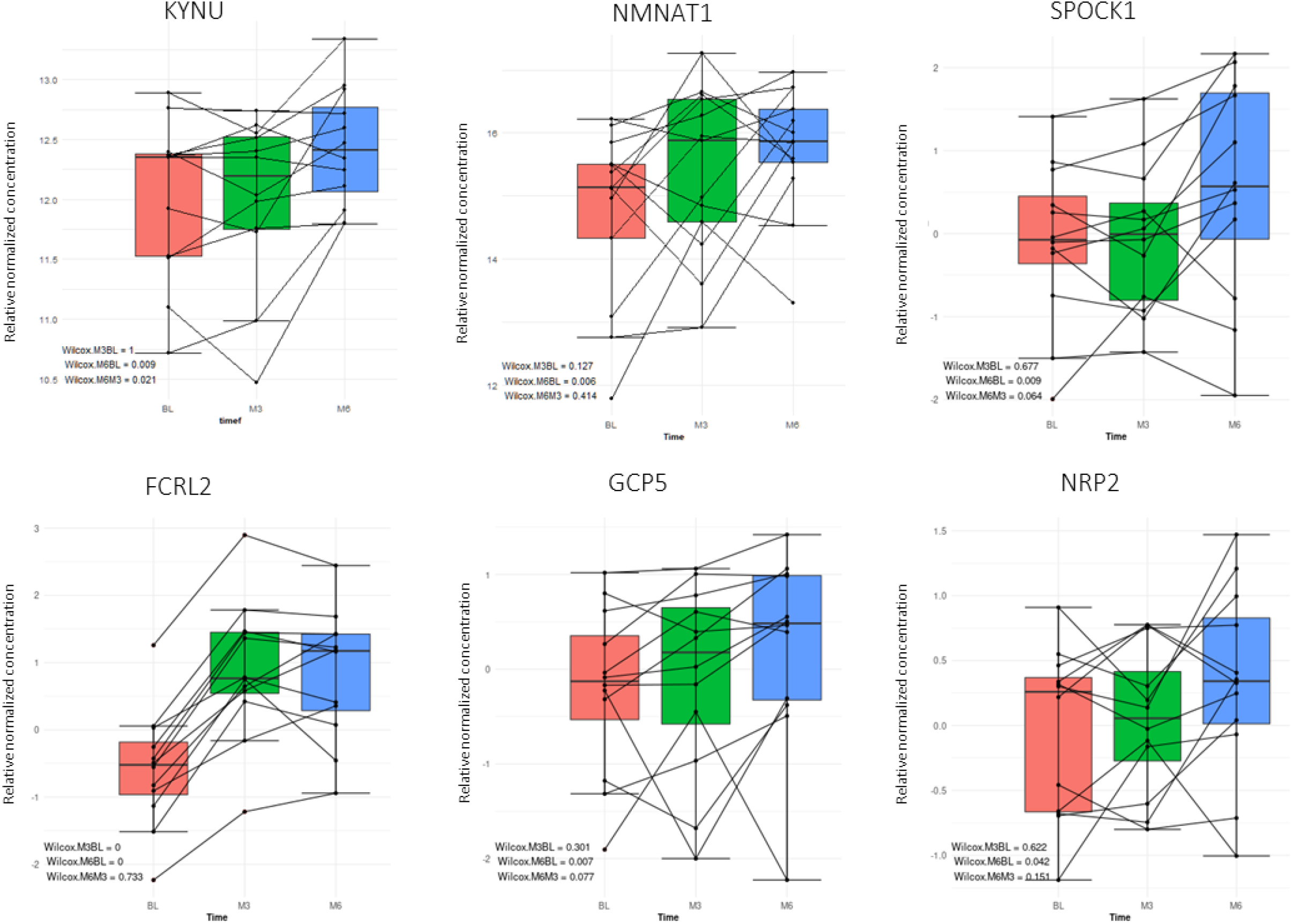
Olink assay analysis of changes in cytokines in 13 patients taking ocrelizumab at baseline (BL), 3 months (M3) and 6 months (M6). Each line represents values for an individual patient at the respective time point in the study. The box plot shows the range (whiskers), first and third quartiles (box), and median (line in box) of the values for each time point. The exact p values shown were calculated using Wilcoxon signed-rank test.

**Figure 5.**
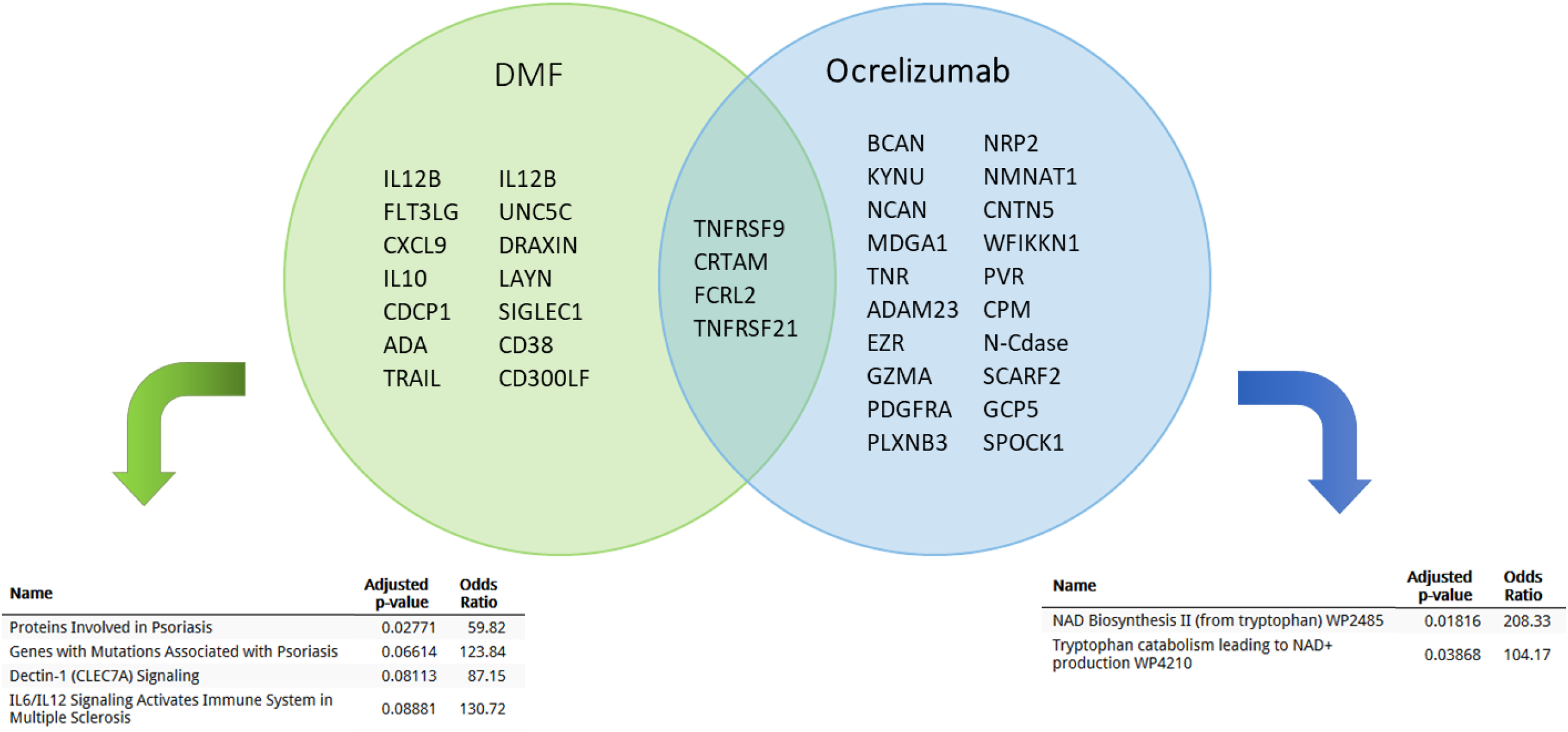
Results of high-throughput, multiplex immunoassay analysis of protein biomarkers in MS patients taking dimethyl fumarate (DMF) and ocrelizumab. Protein biomarkers demonstrating significant (p<0.1) change at month 6 compared to baseline are shown in the Venn diagram. Corresponding gene names were subjected to pathway analysis using EnrichR (https://amp.pharm.mssm.edu/Enrichr/). Top identified pathways (using Elsevier pathway collection for DMF and WikiPathways for ocrelizumab) are listed.

## Discussion

Our study provided high-dimensional lymphocyte and protein profiling of two commonly used MS DMTs. Treatment with DMF led to the downregulation of pro-inflammatory lymphocytes and an associated reduction in inflammatory cytokines. Conversely, B-cell depletion with ocrelizumab resulted in upregulation of neurotropic factors. Immune phenotyping of DMF treated patients confirmed prior findings of modifications to the peripheral immune composition including reductions in memory CD8 and CD4 T cells, memory B cells, and increases in naive and transitional B cells.^2-8, 12-14^ DMF is thought to suppress disease activity in MS by modifying interactions between B and T cells and hindering B cell mediated activation of inflammatory T cells, thereby promoting a more “tolerogenic” immune system.^15^ Our study showed significant reduction in effector T cells including T cell subsets involved in the activation of B cells. Clinical correlations of lower reductions of CD4+, CD8+ T, and B-cells have been shown in DMF treated patients who have stable disease.^8^ Conversely, our study confirms prior findings of increases in circulating transitional and naive B cells.^13^

In contrast with reductions in T and B cells, NK cells have been reported to increase with DMF treatment.^16, 17^ In particular, there was an increase in CD56^bright^ cells, which are known to have regulatory function in MS through cytokine production and lysis of autoreactive T cells.^18^ Upregulation of CD56^bright^ NK cells have also been shown to correlate with reduction in CD8+ memory cells as well as reductions in CD8+ and CD4+ T cells production of inflammatory cytokines,^17^ associating with low disease activity.^15^ These findings overlap with previously established effects of the MS DMT daclizumab, which induces a robust expansion of CD56^bright^ NK cells through selective inhibition of high-affinity IL-2 receptor signaling.^19^

DMF reduced key inflammatory cytokines after 6 months of treatment. CXCL9 is a chemokine that elicits chemotactic activity on T cells upon binding to its receptor CXCR3 expressed in activated Th1 cells. The CXCR3 receptor and its ligands are overexpressed in MS and associated with disease activity.^20, 21^ IL-12 is a proinflammatory cytokine that induces Th1 cell differentiation from naive T cells and stimulate interferon gamma production.^22^ It has been implicated in the pathogenesis of MS, although phase II clinical trial of IL12/23 neutralizing antibody, ustekinumab, did not reduce cumulative gadolinium-enhancing lesions in MS.^23^

We discovered additional new biomarkers potentially decreased by treatment with DMF in our study. SIGLEC1 belongs to the sialic acid binding immunoglobulin-like lectin family and its expression by CD14+ monocytes is increased in MS patients.^24^ In sialoadhesin knockout mice, EAE symptoms were less severe.^25^ Flt3L is a cytokine that plays a role in the mobilization and differentiation of hematopoietic stem cells to polarize toward a Th1 immune response.^26, 27^ Administration of Flt3L have been shown to worsen disease severity in EAE.^28^ TNFRSF9 (CD137) signaling causes activation of microglia and secretion of proinflammatory cytokines.^29^ CD137+ cells were found in MS brain samples with higher concentration of active lesions, while in vitro experiments found CD137 also enhanced inflammatory activity of B cells upon engagement.^30^ Surprisingly anti-inflammatory IL-10 was decreased by DMF treatment, which was also observed in another study of DMT treatment.^13^

Enrichment of corresponding genes of protein biomarkers demonstrating significant (p<0.1) change with 6 months of DMF treatment revealed pathways involved in psoriasis, dectin-1 signaling, and IL-6/IL-12 signaling. Overlap of DMF therapeutic effects with pathways in psoriasis were not surprising considering fumaric acid esters have been used for more than 20 years to treat psoriasis in the EU, and DMF showed efficacy in the BRIDGE trial against placebo in patients with moderate-to-severe chronic plaque psoriasis.^31^ Dectin-1, a C-type lectin, binding to its receptor triggers a signaling cascade leading to the activation of both the canonical and noncanonical the NFkβ pathway.^32^ Dectin-1 deficient mice developed more severe EAE and had mounted increased Th17 responses and had reduced regulatory CD8+ T cells.^33^

Ocrelizumab reduced B cells and increased NK cells. B cells play an important role in antigen presentation and stimulation of T cells and contribute directly to development and progression of MS. In both the periphery and CNS, B cells show signs of antigen-mediated activation through a shift towards antigen-experienced memory B cells.^34^ B cells in MS patients show increased co-stimulatory molecules and increased potential to drive differentiation of inflammatory T cells.^35-37^ Monoclonal antibodies against CD20 deplete B cells but spare plasma cells and hematopoietic stems cells due to lack of CD20 expression. The overlap in effects of DMTs on B cells, in particular memory B cells, suggests the importance role of B cells in the pathogenesis of disease activity in MS. CD20 is also expressed at a low level on a small subset of CD3+ T cells, referred to as CD3+CD20^dim^ T cells.^38^ These CD20+ T cells have been shown to have proinflammatory Th1/Tc1 phenotype and correlate with disease severity in MS.^39^ Anti-CD20 monoclonal antibodies including ocrelizumab and rituximab has also been known to deplete CD3+CD20^dim^ T cells.^40, 41^ Our study showed a trend for a reduction of CD3+CD20^dim^ T cells, but this did not reach statistical significance.

Our data also showed that ocrelizumab treatment induced changes in neurotropic markers related to neural development and axon guidance. The upregulation of neurotropic proteins after 6 months of ocrelizumab treatment suggests a neuroprotective effect of therapy, which we did not observe in our DMF treated participants. The role of B cells in neurodegeneration is not well-understood. Nevertheless, B cells are present in follicle-like aggregates in the meninges of patients with progressive disease course and are associated with microglial activation and neuronal loss.^42^ Differential expression of B-cell specific genes have been associated with high degrees of neurodegeneration, suggesting a role of B-cells in this process.^43^ Our enrichment analysis of significant gene expression affected by ocrelizumab therapy revealed pathways related to NAD biosynthesis and tryptophan catabolism. NAD+ is an important metabolite and cofactor in all living cells, playing a role in processes promoting neuronal resilience and countering the effects of oxidative stress and mitochondrial dysfunction.^44, 45^ Tryptophan catabolism through the kynurenine metabolic pathway produces immunosuppressive metabolites that limit T-cell responses and slowed progression of disease in EAE mice.^46^ In addition, administration of NAD+ has been shown to have neuroprotective effects in promoting myelin and axonal regeneration in EAE.^47^ The mechanistic role of B cell depletion in tryptophan catabolism is not well understood, but may impact indoleamine 2,3-dioxygenase, a key enzyme in regulating humoral immunity that also catalyzes the first and rate limiting step of tryptophan catabolism.^48^

The exploratory nature of our study is limited by the lack of a validation cohort, therefore reproducibility of the study findings remains to be established, especially given the relatively small sample size and the lack of a control group of MS patients not treated with a disease-modifying therapy. In addition, the assay results are subject to increased variation from sample handling due to protein degradation or cell leakage.^49, 50^ Nevertheless, the novel use of proteomic extension technology in our study resulted in the discovery of new biomarkers of treatment response. Reliance on dual-antibody binding and amplification through qPCR provided high assay sensitivity and specificity compared to classical immunoassays for measuring changes in cytokines.

In summary, we identified serum protein biomarkers and associated pathways affected by treatment with DMF and ocrelizumab while confirming the immune profile changes contributing to alterations of immune function. These findings advance our understanding of DMT mechanisms of action and may contribute to identifying additional markers of treatment response or therapeutic targets. Further validation studies are needed to assess these biomarkers’ potential for clinical applications.

## Data Availability

All data produced in the present study are available upon reasonable request to the authors

### Appendix 1. Authors

| Name | Location | Role | Contribution |
| --- | --- | --- | --- |
| Yinan Zhang | Ohio State University, OH | Author | Study design conception; data collection and analysis, drafting and revision of manuscript |
| Brian Lee | Mount Sinai, New York | Author | Data collection, drafting and revision of manuscript |
| Hui Xie | Mount Sinai, New York | Author | Data collection, drafting and revision of manuscript |
| Jonathan Rockoff | Mount Sinai, New York | Author | Data collection, drafting and revision of manuscript |
| Sammita Satyanarayan | Mount Sinai, New York | Author | Drafting and revision of manuscript |
| Rachel Brandstadter | University of Pennsylvania, Pennsylvania | Author | Data collection, drafting and revision of manuscript |
| Seunghee Kim-Schulze | Mount Sinai, New York | Author | Data collection, drafting and revision of manuscript |
| Achillefs Ntranos | Mount Sinai, New York | Author | Study design conception; data collection and analysis, drafting and revision of manuscript |
| Fred Lublin | Mount Sinai, New York | Author | Study design conception, drafting and revision of manuscript |

## References

1. Salmen A and Gold R. Mode of action and clinical studies with fumarates in multiple sclerosis. Exp Neurol. 2014;262 Pt A:52–6.

2. Li R, Rezk A, Ghadiri M, Luessi F, Zipp F, Li H, Giacomini PS, Antel J and Bar-Or A. Dimethyl Fumarate Treatment Mediates an Anti-Inflammatory Shift in B Cell Subsets of Patients with Multiple Sclerosis. J Immunol. 2017;198:691–698.

3. Longbrake EE, Ramsbottom MJ, Cantoni C, Ghezzi L, Cross AH and Piccio L. Dimethyl fumarate selectively reduces memory T cells in multiple sclerosis patients. Mult Scler. 2016;22:1061–1070.

4. Lundy SK, Wu Q, Wang Q, Dowling CA, Taitano SH, Mao G and Mao-Draayer Y. Dimethyl fumarate treatment of relapsing-remitting multiple sclerosis influences B-cell subsets. Neurol Neuroimmunol Neuroinflamm. 2016;3:e211.

5. Staun-Ram E, Najjar E, Volkowich A and Miller A. Dimethyl fumarate as a first-vs second-line therapy in MS: Focus on B cells. Neurol Neuroimmunol Neuroinflamm. 2018;5:e508.

6. Wu Q, Wang Q, Mao G, Dowling CA, Lundy SK and Mao-Draayer Y. Dimethyl Fumarate Selectively Reduces Memory T Cells and Shifts the Balance between Th1/Th17 and Th2 in Multiple Sclerosis Patients. J Immunol. 2017;198:3069–3080.

7. Gross CC, Schulte-Mecklenbeck A, Klinsing S, Posevitz-Fejfar A, Wiendl H and Klotz L. Dimethyl fumarate treatment alters circulating T helper cell subsets in multiple sclerosis. Neurol Neuroimmunol Neuroinflamm. 2016;3:e183.

8. Fleischer V, Friedrich M, Rezk A, Buhler U, Witsch E, Uphaus T, Bittner S, Groppa S, Tackenberg B, Bar-Or A, Zipp F and Luessi F. Treatment response to dimethyl fumarate is characterized by disproportionate CD8+ T cell reduction in MS. Mult Scler. 2018;24:632–641.

9. Montalban X, Hauser SL, Kappos L, Arnold DL, Bar-Or A, Comi G, de Seze J, Giovannoni G, Hartung HP, Hemmer B, Lublin F, Rammohan KW, Selmaj K, Traboulsee A, Sauter A, Masterman D, Fontoura P, Belachew S, Garren H, Mairon N, Chin P, Wolinsky JS and Investigators OC. Ocrelizumab versus Placebo in Primary Progressive Multiple Sclerosis. N Engl J Med. 2017;376:209–220.

10. Assarsson E, Lundberg M, Holmquist G, Bjorkesten J, Thorsen SB, Ekman D, Eriksson A, Rennel Dickens E, Ohlsson S, Edfeldt G, Andersson AC, Lindstedt P, Stenvang J, Gullberg M and Fredriksson S. Homogenous 96-plex PEA immunoassay exhibiting high sensitivity, specificity, and excellent scalability. PLoS One. 2014;9:e95192.

11. Kuleshov MV, Jones MR, Rouillard AD, Fernandez NF, Duan Q, Wang Z, Koplev S, Jenkins SL, Jagodnik KM, Lachmann A, McDermott MG, Monteiro CD, Gundersen GW and Ma’ayan A. Enrichr: a comprehensive gene set enrichment analysis web server 2016 update. Nucleic Acids Res. 2016;44:W90–7.

12. Cunill V, Massot M, Clemente A, Calles C, Andreu V, Nunez V, Lopez-Gomez A, Diaz RM, Jimenez MLR, Pons J, Vives-Bauza C and Ferrer JM. Relapsing-Remitting Multiple Sclerosis Is Characterized by a T Follicular Cell Pro-Inflammatory Shift, Reverted by Dimethyl Fumarate Treatment. Front Immunol. 2018;9:1097.

13. Holm Hansen R, Hojsgaard Chow H, Sellebjerg F and Rode von Essen M. Dimethyl fumarate therapy suppresses B cell responses and follicular helper T cells in relapsing-remitting multiple sclerosis. Mult Scler. 2019;25:1289–1297.

14. Montes Diaz G, Fraussen J, Van Wijmeersch B, Hupperts R and Somers V. Dimethyl fumarate induces a persistent change in the composition of the innate and adaptive immune system in multiple sclerosis patients. Sci Rep. 2018;8:8194.

15. Medina S, Villarrubia N, Sainz de la Maza S, Lifante J, Costa-Frossard L, Roldan E, Picon C, Alvarez-Cermeno JC and Villar LM. Optimal response to dimethyl fumarate associates in MS with a shift from an inflammatory to a tolerogenic blood cell profile. Mult Scler. 2018;24:1317–1327.

16. Marastoni D, Buriani A, Pisani AI, Crescenzo F, Zuco C, Fortinguerra S, Sorrenti V, Marenda B, Romualdi C, Magliozzi R, Monaco S and Calabrese M. Increased NK Cell Count in Multiple Sclerosis Patients Treated With Dimethyl Fumarate: A 2-Year Longitudinal Study. Front Immunol. 2019;10:1666.

17. Smith MD, Calabresi PA and Bhargava P. Dimethyl fumarate treatment alters NK cell function in multiple sclerosis. Eur J Immunol. 2018;48:380–383.

18. Jiang W, Chai NR, Maric D and Bielekova B. Unexpected role for granzyme K in CD56bright NK cell-mediated immunoregulation of multiple sclerosis. J Immunol. 2011;187:781–90.

19. Elkins J, Sheridan J, Amaravadi L, Riester K, Selmaj K, Bielekova B, Parr E and Giovannoni G. CD56(bright) natural killer cells and response to daclizumab HYP in relapsing-remitting MS. Neurol Neuroimmunol Neuroinflamm. 2015;2:e65.

20. Huang J, Khademi M, Fugger L, Lindhe O, Novakova L, Axelsson M, Malmestrom C, Constantinescu C, Lycke J, Piehl F, Olsson T and Kockum I. Inflammation-related plasma and CSF biomarkers for multiple sclerosis. Proc Natl Acad Sci U S A. 2020;117:12952–12960.

21. Simpson J, Rezaie P, Newcombe J, Cuzner ML, Male D and Woodroofe MN. Expression of the beta-chemokine receptors CCR2, CCR3 and CCR5 in multiple sclerosis central nervous system tissue. J Neuroimmunol. 2000;108:192–200.

22. Aslani S, Jafari N, Javan MR, Karami J, Ahmadi M and Jafarnejad M. Epigenetic Modifications and Therapy in Multiple Sclerosis. Neuromolecular Med. 2017;19:11–23.

23. Segal BM, Constantinescu CS, Raychaudhuri A, Kim L, Fidelus-Gort R, Kasper LH and Ustekinumab MSI. Repeated subcutaneous injections of IL12/23 p40 neutralising antibody, ustekinumab, in patients with relapsing-remitting multiple sclerosis: a phase II, double-blind, placebo-controlled, randomised, dose-ranging study. Lancet Neurol. 2008;7:796–804.

24. Malhotra S, Castillo J, Bustamante M, Vidal-Jordana A, Castro Z, Montalban X and Comabella M. SIGLEC1 and SIGLEC7 expression in circulating monocytes of patients with multiple sclerosis. Mult Scler. 2013;19:524–31.

25. Wu C, Rauch U, Korpos E, Song J, Loser K, Crocker PR and Sorokin LM. Sialoadhesin-positive macrophages bind regulatory T cells, negatively controlling their expansion and autoimmune disease progression. J Immunol. 2009;182:6508–16.

26. Maraskovsky E, Brasel K, Teepe M, Roux ER, Lyman SD, Shortman K and McKenna HJ. Dramatic increase in the numbers of functionally mature dendritic cells in Flt3 ligand-treated mice: multiple dendritic cell subpopulations identified. J Exp Med. 1996;184:1953–62.

27. Pulendran B, Banchereau J, Burkeholder S, Kraus E, Guinet E, Chalouni C, Caron D, Maliszewski C, Davoust J, Fay J and Palucka K. Flt3-ligand and granulocyte colony-stimulating factor mobilize distinct human dendritic cell subsets in vivo. J Immunol. 2000;165:566–72.

28. Verda L, Luo K, Kim DA, Bronesky D, Kohm AP, Miller SD, Statkute L, Oyama Y and Burt RK. Effect of hematopoietic growth factors on severity of experimental autoimmune encephalomyelitis. Bone Marrow Transplant. 2006;38:453–60.

29. Yeo YA, Martinez Gomez JM, Croxford JL, Gasser S, Ling EA and Schwarz H. CD137 ligand activated microglia induces oligodendrocyte apoptosis via reactive oxygen species. J Neuroinflammation. 2012;9:173.

30. Wong HY, Prasad A, Gan SU, Chua JJE and Schwarz H. Identification of CD137-Expressing B Cells in Multiple Sclerosis Which Secrete IL-6 Upon Engagement by CD137 Ligand. Front Immunol. 2020;11:571964.

31. Mrowietz U, Szepietowski JC, Loewe R, van de Kerkhof P, Lamarca R, Ocker WG, Tebbs VM and Pau-Charles I. Efficacy and safety of LAS41008 (dimethyl fumarate) in adults with moderate-to-severe chronic plaque psoriasis: a randomized, double-blind, Fumaderm((R)) - and placebo-controlled trial (BRIDGE). Br J Dermatol. 2017;176:615–623.

32. Gringhuis SI, den Dunnen J, Litjens M, van der Vlist M, Wevers B, Bruijns SC and Geijtenbeek TB. Dectin-1 directs T helper cell differentiation by controlling noncanonical NF-kappaB activation through Raf-1 and Syk. Nat Immunol. 2009;10:203–13.

33. Geijtenbeek TB and Gringhuis SI. C-type lectin receptors in the control of T helper cell differentiation. Nat Rev Immunol. 2016;16:433–48.

34. Duddy M, Niino M, Adatia F, Hebert S, Freedman M, Atkins H, Kim HJ and Bar-Or A. Distinct effector cytokine profiles of memory and naive human B cell subsets and implication in multiple sclerosis. J Immunol. 2007;178:6092–9.

35. Aung LL and Balashov KE. Decreased Dicer expression is linked to increased expression of co-stimulatory molecule CD80 on B cells in multiple sclerosis. Mult Scler. 2015;21:1131–8.

36. Genc K, Dona DL and Reder AT. Increased CD80(+) B cells in active multiple sclerosis and reversal by interferon beta-1b therapy. J Clin Invest. 1997;99:2664–71.

37. Harp CT, Ireland S, Davis LS, Remington G, Cassidy B, Cravens PD, Stuve O, Lovett-Racke AE, Eagar TN, Greenberg BM, Racke MK, Cowell LG, Karandikar NJ, Frohman EM and Monson NL. Memory B cells from a subset of treatment-naive relapsing-remitting multiple sclerosis patients elicit CD4(+) T-cell proliferation and IFN-gamma production in response to myelin basic protein and myelin oligodendrocyte glycoprotein. Eur J Immunol. 2010;40:2942–56.

38. Wilk E, Witte T, Marquardt N, Horvath T, Kalippke K, Scholz K, Wilke N, Schmidt RE and Jacobs R. Depletion of functionally active CD20+ T cells by rituximab treatment. Arthritis Rheum. 2009;60:3563–71.

39. von Essen MR, Ammitzboll C, Hansen RH, Petersen ERS, McWilliam O, Marquart HV, Damm P and Sellebjerg F. Proinflammatory CD20+ T cells in the pathogenesis of multiple sclerosis. Brain. 2019;142:120–132.

40. Gingele S, Jacobus TL, Konen FF, Hummert MW, Suhs KW, Schwenkenbecher P, Ahlbrecht J, Mohn N, Muschen LH, Bonig L, Alvermann S, Schmidt RE, Stangel M, Jacobs R and Skripuletz T. Ocrelizumab Depletes CD20(+) T Cells in Multiple Sclerosis Patients. Cells. 2018;8.

41. Palanichamy A, Jahn S, Nickles D, Derstine M, Abounasr A, Hauser SL, Baranzini SE, Leppert D and von Budingen HC. Rituximab efficiently depletes increased CD20-expressing T cells in multiple sclerosis patients. J Immunol. 2014;193:580–586.

42. Magliozzi R, Howell O, Vora A, Serafini B, Nicholas R, Puopolo M, Reynolds R and Aloisi F. Meningeal B-cell follicles in secondary progressive multiple sclerosis associate with early onset of disease and severe cortical pathology. Brain. 2007;130:1089–104.

43. Comabella M, Canto E, Nurtdinov R, Rio J, Villar LM, Picon C, Castillo J, Fissolo N, Aymerich X, Auger C, Rovira A and Montalban X. MRI phenotypes with high neurodegeneration are associated with peripheral blood B-cell changes. Hum Mol Genet. 2016;25:308–16.

44. Lautrup S, Sinclair DA, Mattson MP and Fang EF. NAD(+) in Brain Aging and Neurodegenerative Disorders. Cell Metab. 2019;30:630–655.

45. Pehar M, Harlan BA, Killoy KM and Vargas MR. Nicotinamide Adenine Dinucleotide Metabolism and Neurodegeneration. Antioxid Redox Signal. 2018;28:1652–1668.

46. Platten M, Ho PP, Youssef S, Fontoura P, Garren H, Hur EM, Gupta R, Lee LY, Kidd BA, Robinson WH, Sobel RA, Selley ML and Steinman L. Treatment of autoimmune neuroinflammation with a synthetic tryptophan metabolite. Science. 2005;310:850–5.

47. Tullius SG, Biefer HR, Li S, Trachtenberg AJ, Edtinger K, Quante M, Krenzien F, Uehara H, Yang X, Kissick HT, Kuo WP, Ghiran I, de la Fuente MA, Arredouani MS, Camacho V, Tigges JC, Toxavidis V, El Fatimy R, Smith BD, Vasudevan A and ElKhal A. NAD+ protects against EAE by regulating CD4+ T-cell differentiation. Nat Commun. 2014;5:5101.

48. Shinde R, Shimoda M, Chaudhary K, Liu H, Mohamed E, Bradley J, Kandala S, Li X, Liu K and McGaha TL. B Cell-Intrinsic IDO1 Regulates Humoral Immunity to T Cell-Independent Antigens. J Immunol. 2015;195:2374–82.

49. Shen Q, Bjorkesten J, Galli J, Ekman D, Broberg J, Nordberg N, Tillander A, Kamali-Moghaddam M, Tybring G and Landegren U. Strong impact on plasma protein profiles by precentrifugation delay but not by repeated freeze-thaw cycles, as analyzed using multiplex proximity extension assays. Clin Chem Lab Med. 2018;56:582–594.

50. Tworoger SS and Hankinson SE. Collection, processing, and storage of biological samples in epidemiologic studies: sex hormones, carotenoids, inflammatory markers, and proteomics as examples. Cancer Epidemiol Biomarkers Prev. 2006;15:1578–81.

